# Developing a training on disability for healthcare workers in Uganda: a mixed methods study

**DOI:** 10.1101/2023.07.25.23293148

**Authors:** Tracey Smythe, Andrew Sentoogo Ssemata, Sande Slivesteri, Femke Bannink Mbazzi, Hannah Kuper

## Abstract

**Background:** Approximately 1.3 billion people worldwide face barriers in accessing inclusive healthcare due to disabilities, leading to worse health outcomes, particularly in low and middle-income countries (LMIC). However, there is a lack of training of healthcare workers about disability, both globally and in Uganda.

**Objectives:** To use mixed methods to develop a comprehensive training program with standardised elements for healthcare workers in Uganda, focusing on improving their knowledge, attitudes, and skills in providing care for people with disabilities.

**Methods:** The Medical Research Council (MRC) approach was employed to guide the development of the training intervention. We conducted an umbrella review to gather relevant literature on disability training for healthcare workers. Interviews were conducted with international experts to gain insights and perspectives on the topic. Additionally, interviews were undertaken with people with disabilities and healthcare workers in Uganda to understand their experiences and needs. A participatory workshop was organised involving key stakeholders, to collaboratively design the training material based on the findings from these data sources.

**Results:** Eight review articles examined training programs for healthcare workers on disability. Training settings ranged from specialised clinical settings to non-clinical settings, and the duration and evaluation methods of the training varied widely. Lectures and didactic methods were commonly used, often combined with other approaches such as case studies and simulations. The impact of the training was assessed through healthcare worker reports on attitudes, knowledge, and self-efficacy. Interviews emphasised the importance of involving people with disabilities in the training and improving communication and understanding between healthcare providers and people with disabilities. Five themes for healthcare worker training on disability were generated through the workshop, including responsibilities and rights, communication, informed consent, accommodation, and referral and connection, which were used to guide the development of the curriculum, training materials and training approach

**Conclusions:** This study presents a novel approach to develop a training program that aims to enhance healthcare services for people with disabilities in Uganda. The findings offer practical insights for the development of similar programs in LMICs. The effectiveness of the training program will be evaluated through a pilot test, and policy support is crucial for its successful implementation at scale.

**Key messages:**

1. Healthcare workers require training to effectively address the health concerns of people with disabilities, yet this is rarely included in curricula worldwide
2. Uganda recognises the importance of addressing this issue and is taking steps to improve training programs about disability for healthcare workers
3. We used mixed methods to co-develop a comprehensive training program with standardised elements for healthcare workers in Uganda, focusing on improving their knowledge, attitudes, and skills in providing care for people with disabilities.
4. The developed training material could be adapted for healthcare workers in other resource-limited settings, and policy support is needed to ensure its implementation at scale

## Introduction

Approximately 16% of the world’s population, or 1.3 billion people, live with a disability, and the majority live in low and middle income countries (LMIC) (1). Access to inclusive healthcare services is vital for promoting health equity and social inclusion for all (2). However, healthcare workers’ lack of knowledge, skills, and attitudes towards disability remains a significant barrier to achieving this goal (3). Efforts to integrate disability-related education and training into healthcare curricula and continuing education programs, as emphasized by the WHO report (1), are crucial in addressing this issue. Equipping healthcare workers with the necessary tools and resources to provide effective, respectful, and culturally sensitive care to people with disabilities is essential in meeting their needs and promoting positive health outcomes. People with disabilities want to be “expected, accepted and connected” by the health system (2). This requires training to ensure that healthcare staff are aware of disabilities, possess relevant skills, and maintain positive attitudes, enabling them to make appropriate linkages to other necessary care. Efforts to integrate disability-related education and training into healthcare curricula and continuing education programs are crucial in addressing this issue (1, 2, 4).

Training on disability can lead to improvements in the knowledge and attitudes of healthcare workers towards people with disabilities throughout all stages of their careers. For example, two medical colleges in the US integrated disability across medical student training curricula, and medical students improved their knowledge, attitudes, and core competencies in treating patients with disabilities (5). In Rwanda, continuing professional development training about childhood disability used case studies and clinic visits, and instructors gave participants’ immediate feedback on their practice. As a result, participants demonstrated improved clinical decision making skills in paediatric rehabilitation (6). Similarly, programmes that invited people with disabilities as teachers found that participants believed the nonclinical interaction enhanced their comfort and attitudes towards people with disabilities (7, 8) Despite the global recognition that well-trained, fairly distributed and motivated healthcare workers are critical to improving population health and achieving universal health coverage (UHC) and Sustainable Development Goals (SDGs) (9), there is a lack of standardised programs focusing on disability (10), especially in LMICs. Existing programs covering diverse content areas such as general disability awareness, condition-specific knowledge, rehabilitation skills, assistive technology, inclusive design, universal design for learning, mental health, and primary healthcare (Table 1). There has been little consideration to date on what is optimal in terms of content of disability training for healthcare workers.

**Table 1:**
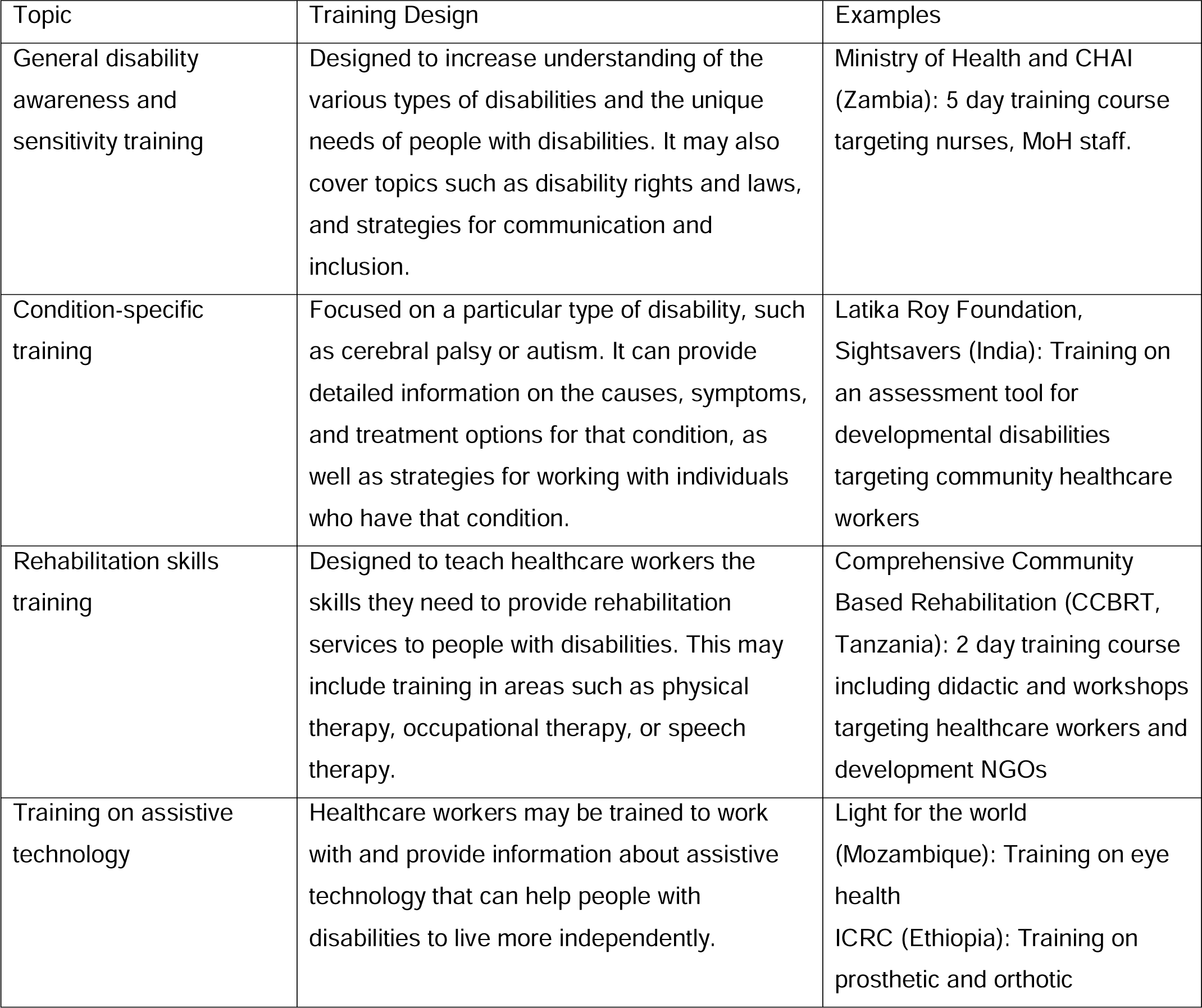
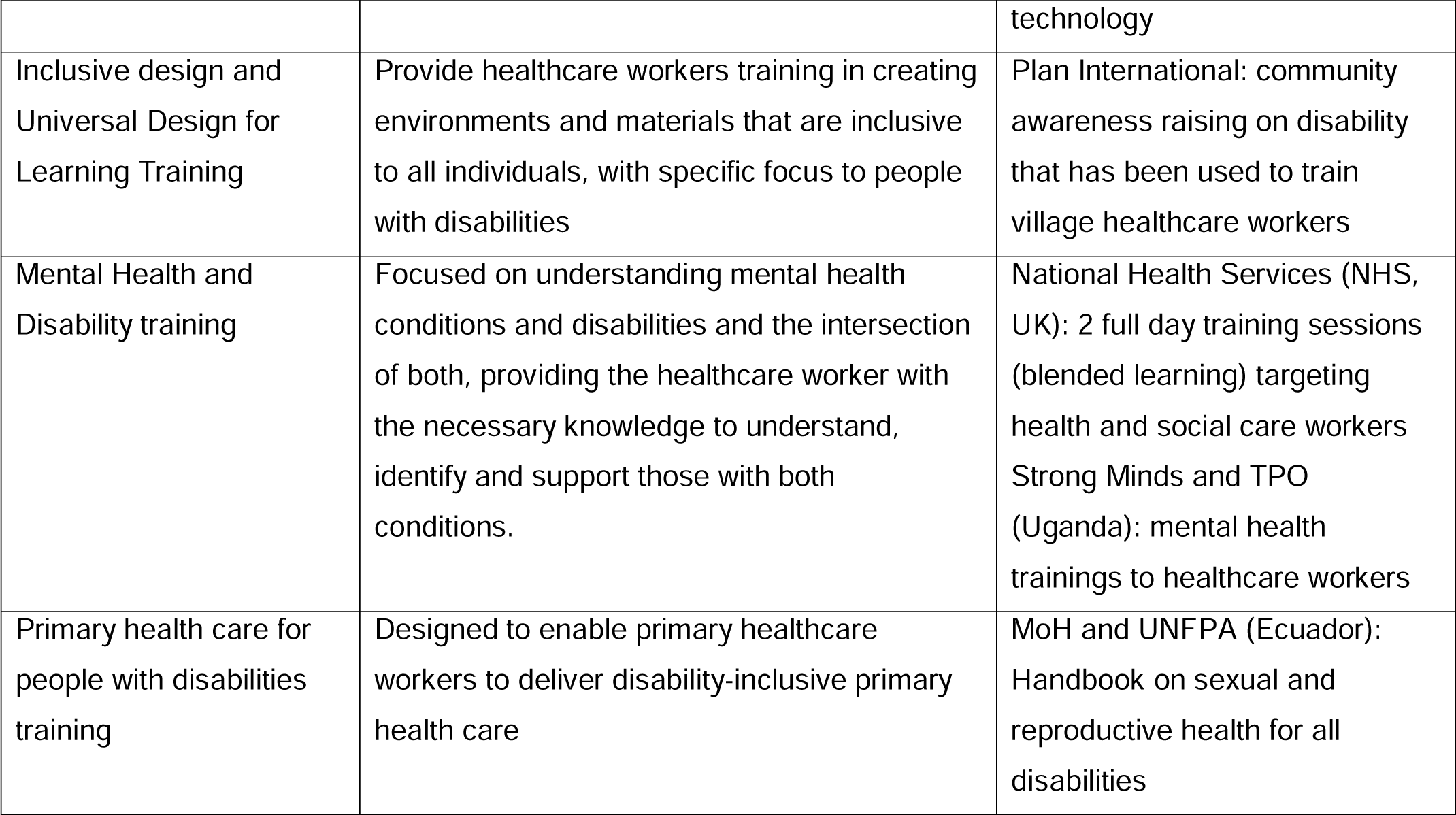
Examples of training programmes delivered to healthcare workers on disability.

There are various potential approaches to training, including face-to-face or remote, involvement of people with disabilities or not, and emphasis on knowledge, skills, or attitudes, as well as different underlying models, such as medical or rights-based (10, 11). Despite this range of possibility, there is limited collation or scrutiny of information, including input from people with disabilities and healthcare workers, to identify the most effective approach. Additionally, LMICs face unique challenges in healthcare provision, including resource constraints and limited access to education and training (12), which may contribute to the acute lack of training opportunities for healthcare workers. For instance, Uganda encounters significant challenges in training healthcare workers on disability due to limited resources, infrastructure, and cultural barriers that may stigmatise or exclude people with disabilities (13–15).

The objective of this study was therefore to use mixed methods to develop a comprehensive training program with standardised elements for healthcare workers in Uganda, focusing on improving their knowledge, attitudes, and skills in providing care for people with disabilities. This paper describes the development of the training and presents the training material developed. We provide an in-depth examination of the training needs of healthcare workers in the area of disability and offer a practical solution in the form of the developed training material that can be used to improve the knowledge and skills of healthcare workers on disability. A future study will pilot test and evaluate the training material.

## Methods

We used the Medical Research Council (MRC) approach, which involves a systematic and evidence-based framework to development of a programme (16). It emphasises the importance of involving stakeholders throughout the process and ensures that interventions are evidence-based, feasible, and acceptable to those involved in their implementation. This approach aims to create effective and sustainable interventions that improve health outcomes and services.

### Overview of methods

This study utilised data from an umbrella review, interviews with international experts on disability training for healthcare workers, and interviews with people with disabilities and healthcare workers in Uganda. The data informed the design of a workshop that involved key stakeholders, including healthcare workers and people with disabilities, in co-creating the training material. The training material was developed based on information gathered from the umbrella review, interviews, and design workshop.

### Umbrella review

The umbrella review was conducted to identify systematic reviews and meta-analyses of studies that examined associations between training of healthcare workers on any disability and change in healthcare worker behaviour, attitude or treatment delivered. The protocol for this study was registered in the International Prospective Register of Systematic Reviews (PROSPERO), reference number #CRD42023390881. We used the Preferred Reporting Items for Overviews of Reviews (PRIOR) statement for conducting umbrella reviews. We searched PubMed for studies published in English between 1^st^ January 2015 and 11^th^ January 2023, using the terms (“train*” OR “educat*”) AND (“healthcare worker” OR “health professional” OR “medical professional”) AND (“disability” OR “impairment”) filtered for systematic reviews and meta-analyses for this rapid umbrella review. Inclusion criteria were established as: systematic reviews or meta-analyses that evaluated training on disability (intervention) for any health professional (population). Studies were excluded if published prior to 2015 to ensure the currency and relevance of information, and if in any other language than English. We searched reference lists of included studies for additional eligible papers.

Two reviewers (TS, ASS) independently assessed study eligibility and extracted the data. The risk of bias of the included studies was assessed using AMSTAR 2 (A MeaSurement Tool to Assess systematic Reviews) (17), designed to appraise systematic reviews that include randomised controlled trials. The instrument provides a broad assessment of quality, including flaws that may have arisen through poor conduct of the review with uncertain impact on findings. We developed and pilot-tested an extraction tool in Excel, to systematically record information from included studies. Extracted information included: 1) Publication characteristics: author, title, year of publication, setting/country; 2) Study design: study design, sample size; 3) Participant characteristics: cadre, and any other relevant descriptive data; 4) Outcomes: effect size of training. Data were extracted by TS and checked for accuracy by ASS. Where studies included information on training other professionals (e.g. police officers, teachers) only data on healthcare workers were extracted. Data were also only extracted on training when reviews included additional information (e.g use of disability measurement tools). We narratively synthesised the results.

### Semi-structured interviews

Interviews were conducted with international experts with experience of delivering training on disability in January 2022, in order to explore current practices and to identify gaps in practice and policy. Six experts were purposively sampled to represent people with and without disabilities, in low and high income settings. They were interviewed by TS, a physiotherapist and epidemiologist from Zimbabwe with mixed-methods expertise. Interviews were held online using Zoom. Verbal informed consent was given. A set of open-ended questions (Supplementary file 1) elicited detailed responses on the experiences, perspectives and practices of experts in the field of healthcare worker training on disability. The interviews were recorded and transcribed for analysis. Transcripts were coded and thematically analysed using NVIVO.

Interviews were then conducted in Uganda with people with disabilities and healthcare workers. Written consent was given. People with disabilities were asked what they wished healthcare workers would know about disability, and interviews with a range of healthcare workers were used to gather more detailed information on their specific training needs and to understand their perceptions of the current training available Semi-structured interviews were conducted in-person with 17 healthcare workers and 27 people with disabilities in Luuka District, Uganda. ASS and SS, Ugandan social scientists with expertise in qualitative methodology undertook the interviews. Participants were recruited through existing networks, non-government organisations and health facilities. The healthcare workers were selected from the eight sub-counties that make up Luuka district based on cadre (Clinicians, Midwives, Nurses, counsellors, laboratory technicians), and level of health facility (health centre, II,III or IV) while the people with disabilities were purposively selected to ensure representation of age, gender and impairment. The interviews explored experiences and perspectives of delivering and receiving health services (Supplementary file 2: Healthcare worker, Supplementary file 3: people with disability). The interviews were conducted in a private and comfortable setting and were audio recorded and transcribed. The transcripts were coded and manually analysed using a thematic analysis approach.

### Design workshop

A design workshop was held to develop a framework for healthcare worker training on disability in Entebbe Uganda in September 2022. The workshop brought together a group of people with diverse disabilities (n=7) and healthcare workers and medical educationalist (n=5) to actively participate in the design process over 2 days. The workshop was facilitated by TS and ASS. The workshop consisted of a series of participatory activities, including group discussions, brainstorming sessions, and small group exercises. These activities were designed to elicit the perspectives, experiences, and recommendations of participants. The workshop concluded with a consensus-building activity, where participants discussed and agreed on the key components of the training framework.

### Development of training material

A theory of change was used to inform the content design and implementation of the training framework. The research team created the first theory of change model, led by TS, drawing on information gathered from the international experts. The theory of change approach involved identifying the desired outcomes of the training and the necessary steps and activities to be put in place to achieve those outcomes. The training material was then developed by the study team drawing on evidence from the umbrella review, interviews and design workshop on the most effective training strategies for healthcare workers on disability, and was refined based on feedback from five people with disabilities.

## Results

The literature search for the umbrella review yielded 377 studies, with 24 full-text articles selected for further review. Eight review articles were eligible for inclusion in the final analysis (Fig 1). A total of 227 studies were included in the reviews, but only 4 studies were conducted in LMICs. The reviews related to training in intellectual and developmental disabilities, mental health, and all disabilities. Three systematic reviews were rated with high confidence on the AMSTAR2 tool, two rated with medium and three rated with low confidence (supplementary file 4). The systematic reviews receiving a low confidence rating in their findings were evaluated as such because they failed to pre-register the review and did not include an appropriate evaluation of bias, including publication bias of the included studies.

**Fig 1:**
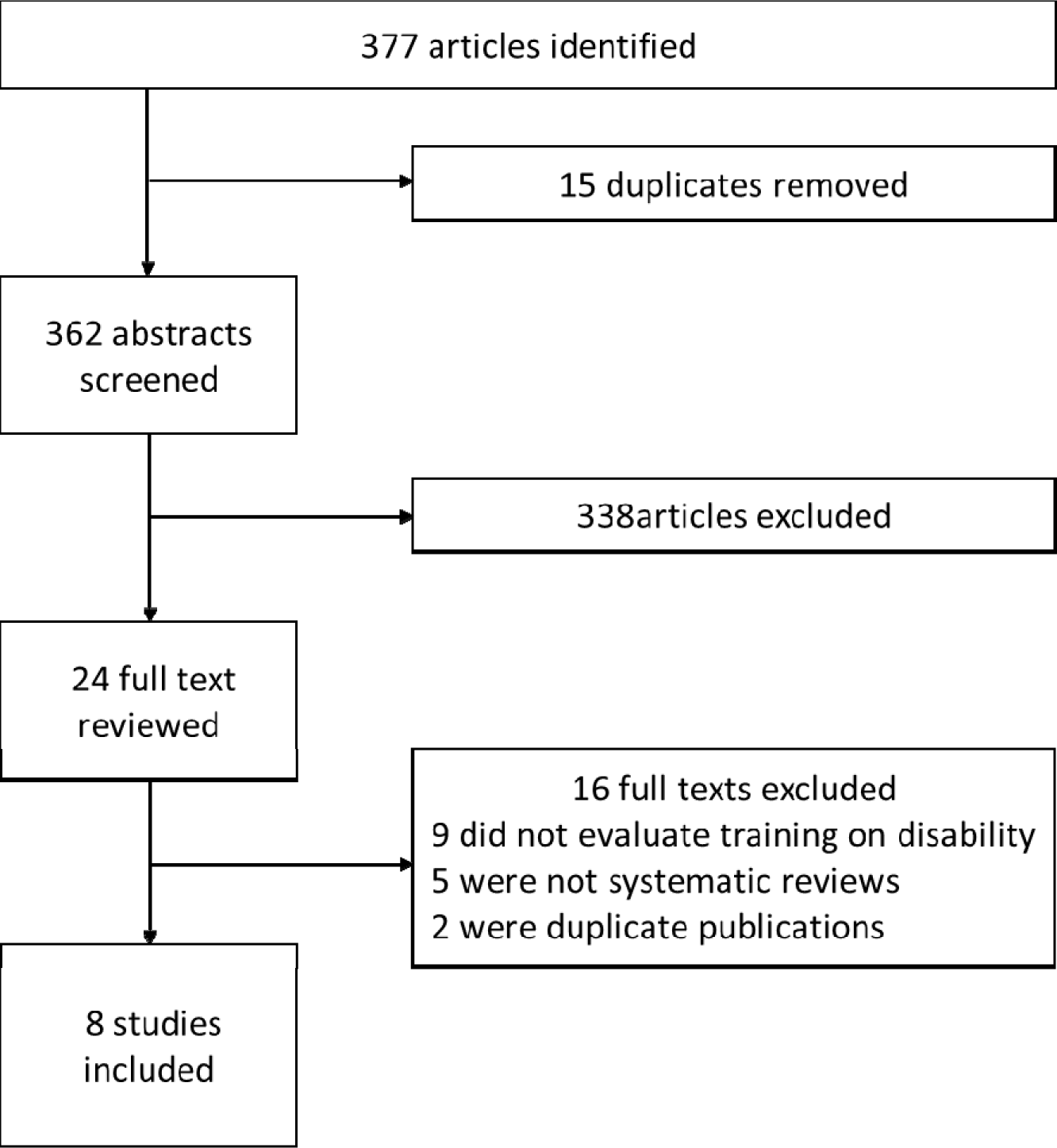
PRISMA for umbrella review of systematic reviews and meta-analyses on training of healthcare workers on disability.

The training settings included specialised clinical settings, non-specialised clinical settings (inpatient and outpatient), continuity-clinic based, non-clinical settings, medical schools, GP practices, primary care clinics, and various other settings such as clinics, camps, schools, residential settings, and universities. The timeline of training and training evaluation methods varied across the articles; Some articles reported single-session training, short-term training (<1 month), and longer-term training (1-3 months or >3 months).

Lectures and didactic (instructional) methods were the most commonly reported teaching methods in disability education, often combined with other approaches, such as case studies, simulations, and placements. Some training programmes leverage multimedia tools, such as video recording or simulations, to enhance the learning experience. The content of the education typically includes disability from a rights-based perspective, as well as particular skills, such as sign language for medical and pharmacology terms (Table 2). Many programmes involved people with disabilities as active participants in the education, such as simulating patients or serving as teachers to help run activities. Through these interactions, contact with people with disabilities was transformative, leading to significant changes in attitudes and perceptions of participants.

A broad variety of evaluation methods were used, such as pre- and post-test knowledge assessments, questionnaires at baseline up to 18 months, and immediate post-training evaluations. The impact of the education was typically measured by healthcare worker reports of comfort and attitudes towards people with disabilities, as well as communication skills, knowledge, and self-efficacy. Target outcomes for the training interventions varied, with some focusing on perspective/awareness/comfort, medical and clinical knowledge. Several had unclear outcomes. However, only a few studies considered the longer-term impact, specifically three months post-intervention.

**Table 2:**
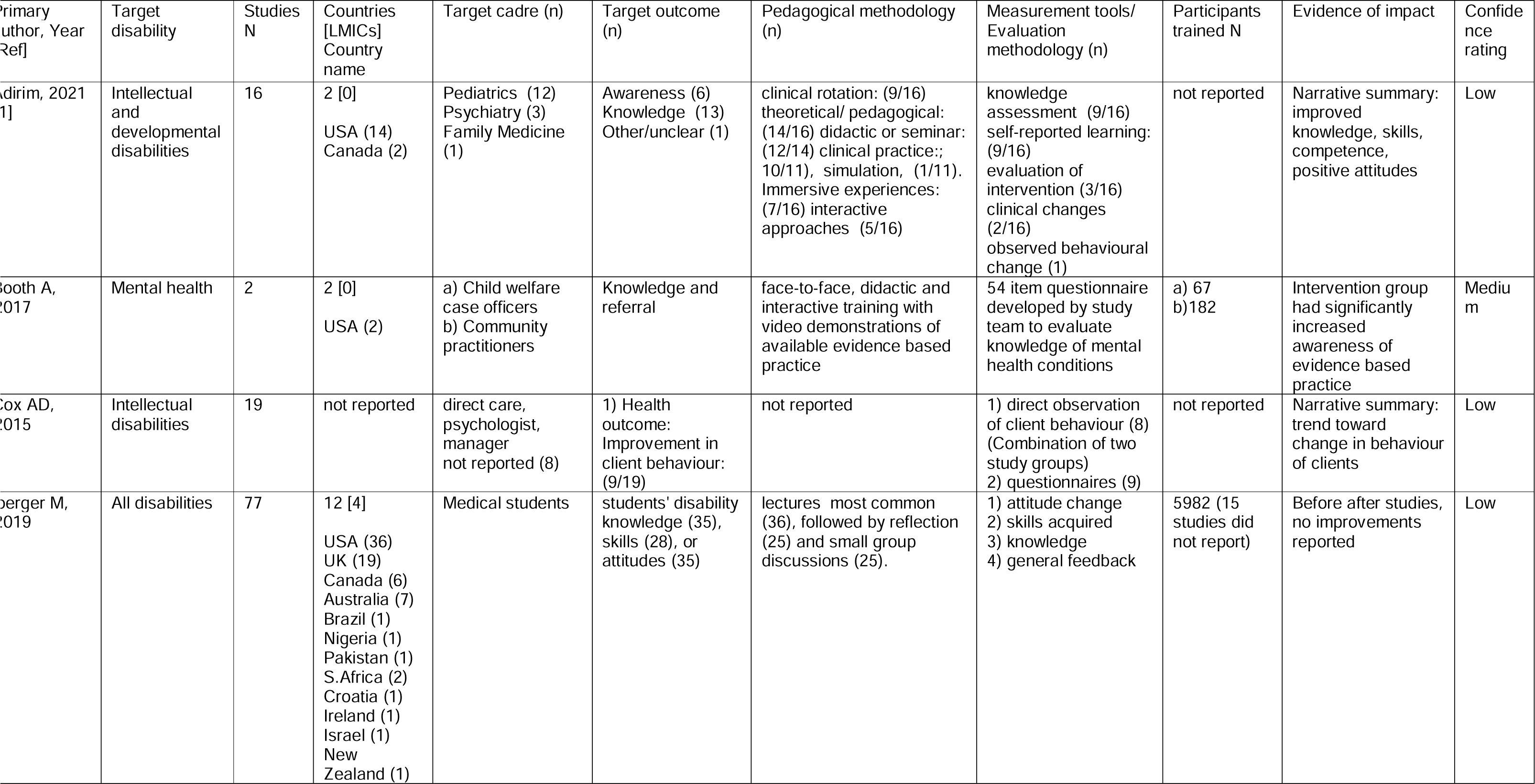
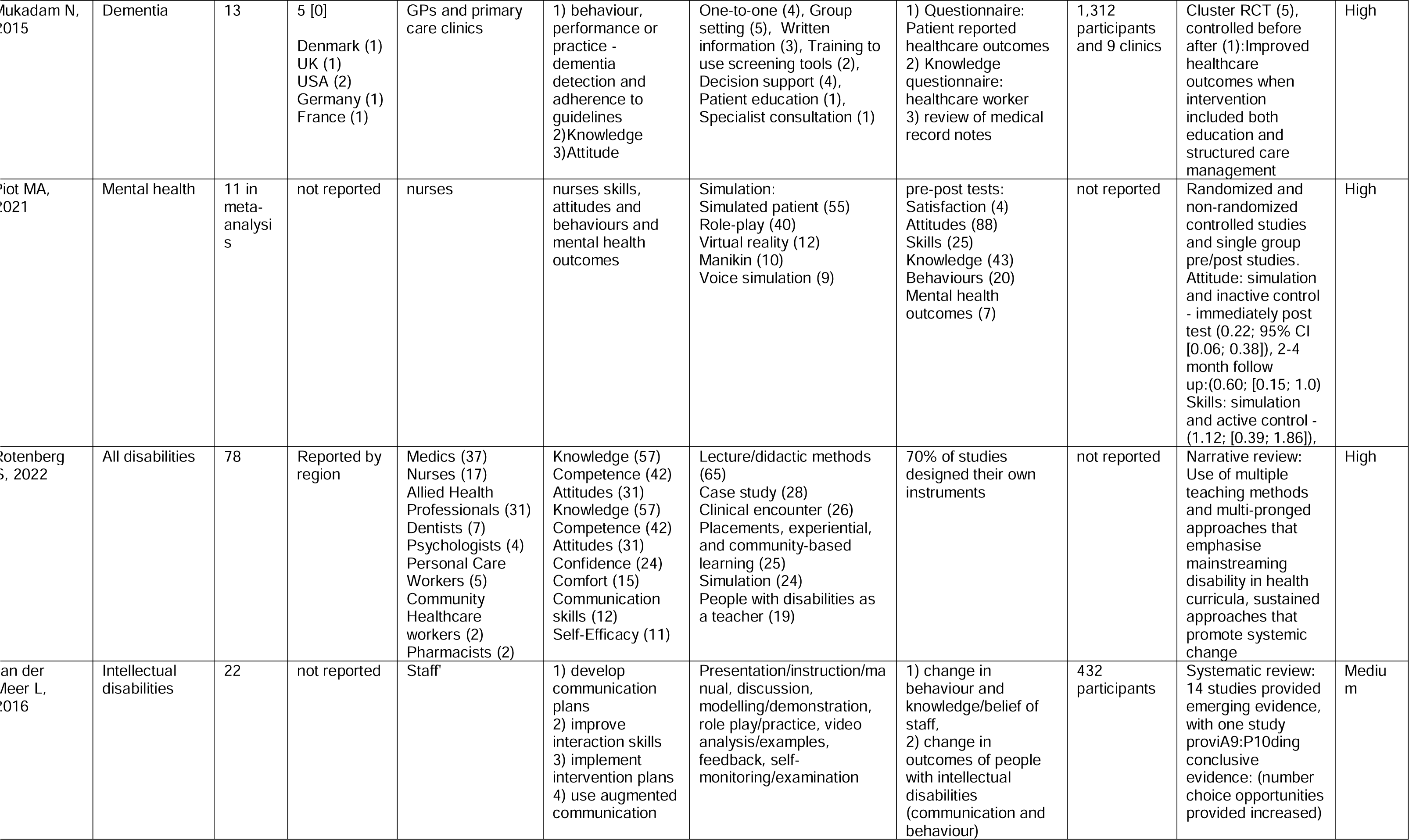
Training on disability effect estimates for healthcare workers reported in systematic reviews (2015–2022)

### Interviews with international experts

International experts reported that people with disabilities play an important role in improving the quality of care for themselves and others. Interviewees identified several ways that this role would occur, as people with disabilities can: offer unique insights into their experiences and perspectives, identify areas for improvement in their care, serve as advocates and role models, and help to promote a culture of inclusion and understanding in the healthcare system. They can also help to ensure that training programs are relevant, effective, and responsive to the needs of people with disabilities.

Experts reported that good practice examples of training methods included contextualized story-telling and activities adapted by the trainer to the local context. Participants believed that these methods were most effective in engaging learners and improving their knowledge, attitudes, and skills related to disability. However, the success of these methods depended on the quality of the training materials, the experience and skills of the trainers, and the level of buy-in and engagement from learners.

Interviewees identified challenges in providing disability education within healthcare training, citing issues such as lack of standardization in curricula and limited time and resources. They expressed concern that these factors may contribute to inadequate understanding of disability issues among healthcare providers, resulting in disparities in care and outcomes for people with disabilities. Additionally, participants discussed emerging trends in healthcare, such as the use of telemedicine and wearable devices, which they believed would require healthcare workers to develop an even deeper understanding of the needs of people with disabilities. They identified opportunities for scaling training, including integrating disability education into undergraduate degrees, continuing education programs, mentoring, and coaching. Overall, participants viewed the future of disability education for healthcare workers as an exciting opportunity for growth and advancement in the healthcare industry.

### Interviews in Uganda

Overall, the importance of improving communication, understanding, and collaboration between healthcare providers and people with disabilities to promote equitable and high-quality healthcare services was emphasised in interviews with both people with disabilities and healthcare workers.

People with disabilities highlighted that they expected healthcare workers to recognise and be made aware of the challenges they face when trying to access healthcare services. Additionally, they emphasised that they desired to be treated with the same respect and dignity as any other patient, and they value a positive attitude from healthcare providers towards their care.

> *“We want healthcare workers to know that all people are equal, including those of us with disabilities, and they should treat each person with dignity.”* (Male, visual impairment)
>
> *“The doctors in the hospital should know that we too are humans and have blood like them but the difference maybe is that one of my parts is weak but it doesn’t stop us from getting sick like them.”* (Female, physical impairment)

Many of the participants acknowledged that better communication skills from healthcare providers, such as clear and concise explanations, are essential to building trust and rapport with people with disabilities.

> *“What I know is that for my life to continue being good, it is important for me to be able to visit a healthcare worker who understands my situation and I am not insulted…The care and respect you give me when you see me and you speak with is helpful.”* (Male, Albinism)

Similarly, the healthcare workers expressed a need for more training and education on disability to provide high-quality care. The reported training needs ranged from the basic training orientation on disability, including communication skills, knowledge on how to navigate the time needed for disability-inclusive care, and how to examine and treat people with disabilities during routine health visits.

> *“First of all, we need training because people with disabilities cannot be managed like other individuals. We need training on the forms of disabilities because the different types of disabilities call for different management. So, we can be empowered …and we can manage the expected and non-expected challenges that these people can have.”(Male, Medical officer)*
>
> *“We need to know, how do you assess them, how do you support them and counsel them. If you do not have those skills sometimes you can mishandle them. For example, you may just look at the cough they have but behind the cough there could be other things.”* (Female, Senior nursing officer)

Furthermore, healthcare providers pointed out the need for adequate information about specialised service needs, how to make referrals and contextual considerations (e.g. cultural, social, economic) to ensure that people with disabilities received appropriate care and were referred for further management.

> *“If I recognise the need for specialised care, I would simply write a referral note. However, many times I am unsure of where to refer them, so I am unable to follow up on whether they received the necessary assistance. Writing a referral is the best I can do.”* (Male, Senior medical officer)
>
> *“We can train healthcare workers within the facility on the best practices for interacting with people with disabilities. Even those of us who support them in the community can be trained on how to connect them with specialised services*.” (Male, VHT coordinator)
>
> *“It may be beneficial to collaborate with others who work with individuals with disabilities and provide holistic care. Since we operate at different levels and some hospitals have specialised clinics for individuals with disabilities, working together can help ensure they receive proper services”.* (Female, Midwife)

Involving individuals with disabilities in health workshops was regarded as a way of facilitating mutual understanding and enabling healthcare workers to better address the specific needs of persons with disabilities, while establishing a designated person for follow-up can enhance continuity of care.

> *“Sometimes we are left behind, but if we are invited to health workshops, we can share our experiences with healthcare workers, including how we feel and how we should be treated. We can discuss the specific disabilities we face and the challenges we encounter. This would provide an opportunity for healthcare workers to better understand and appreciate what individuals with disabilities go through.*” (Male, Person with disability councillor, Visual impairment)

The findings suggest that healthcare workers often feel uncertain about referring individuals with disabilities for specialised care, leading to challenges in ensuring necessary and appropriate healthcare. Training healthcare workers on best practices for interacting with people with disabilities, both within the facility and in the community was recommended to help improve their ability to connect individuals with specialised services and provide holistic care.

### Themes and recommendations from the participatory workshop

Five themes for healthcare worker training on disability were generated by consensus from discussions during the participatory workshop. These themes include the need for a focus on:

- responsibilities and rights: emphasising understanding of the rights of people with disabilities
- communication: highlighting effective communication for building rapport
- informed consent: focusing on respecting privacy during examinations
- accommodation: advocating for disability-inclusive practices
- referral and connection: ensuring appropriate referrals and connections to other healthcare services.

The workshop also emphasised the importance of active participation, clear communication, and inclusivity in the design process.

It was recommended that the training program should adopt a modular approach, focusing on different aspects of providing healthcare to people with disabilities. It should also focus on increasing knowledge and understanding of disabilities, improving the attitudes and practices of healthcare workers towards people with disabilities. Emphasis should be placed on adopting a disability-inclusive approach, training healthcare workers on the social model of disability, recognising the impact of societal barriers on the lives of people with disabilities, and addressing these barriers to promote greater inclusion and participation in society. Cultural sensitivity and the use of appropriate terminology are essential help to ensure that the training is inclusive and relevant to the diverse population of people with disabilities that healthcare workers may encounter.

Interaction with people with disabilities was recommended as a key component. This will involve inviting people with disabilities to participate in the training sessions and share their experiences and perspectives. The training should also adopt a learner-centred and participatory approach, based on the values of the healthcare worker, promoting reflection on their own values and beliefs and applying them in their practice, which may be effective in promoting a sense of ownership and commitment to working with people with disabilities.

Ongoing mentorship and peer learning should support the training, pairing healthcare workers with experienced mentors to provide guidance and support as they apply their new knowledge and skills in practice. Practical skills should be included in the training, such as techniques for measuring the weight of a person with a physical impairment, to ensure comprehensive and considerate examination and treatment of people with disabilities.

### Developed training material

The theory of change provided a clear and logical structure for the development of the training framework and helped to ensure that the framework was comprehensive, evidence-based and responsive to the specific needs and experiences of people with disabilities in Uganda. The desired outcomes of the training included increasing the knowledge, skills and attitudes of healthcare workers on disability, to ensure that people with disabilities are expected, accepted and connected within health systems. The ceiling of responsibility is the distal outcome that healthcare workers provide disability inclusive care that is considerate and comprehensive. Key components and activities that would be necessary to achieve the proximal outcomes included interaction with people with disabilities, a learner centred participatory approach based on the values of healthcare workers, ongoing mentorship and peer learning with an African health disability perspective (Fig 2).

**Figure 2:**
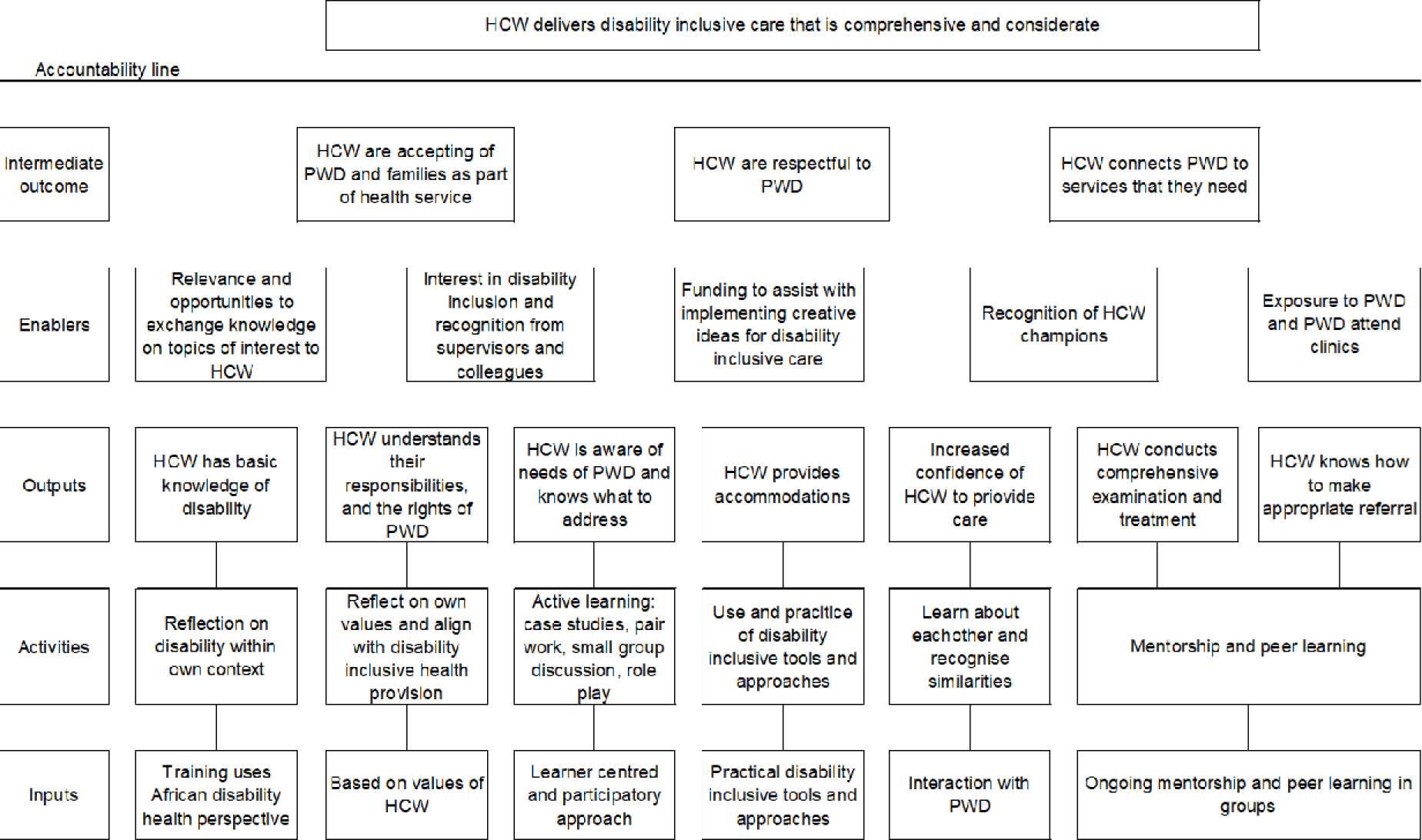
Theory of change. *HCW = healthcare worker, PWD = people with disabilities

The developed training programme included a range of activities such as pre-training self-assessments, interactive workshops, case studies and mentoring. The training framework also included specific provisions for accessibility and inclusion, recognising the importance of addressing the specific needs and experiences of people with disabilities.

The training will be delivered by a healthcare worker and a person with a disability. They will facilitate the training in person over one day for various healthcare workers (nurses, technicians, community health workers, allied health professionals, doctors). The programme will focus on key areas such understanding disability, good practices in work and context, and personal motivations for providing disability-inclusive healthcare. Participants will learn about routine health needs, communication skills, assessing and treating persons with disabilities, appropriate referrals, and ensuring accessibility in healthcare settings.

The training program aims to achieve its goals through several strategies. It promotes a disability-inclusive approach by engaging people with disabilities and encouraging active participation and reflection. Ongoing mentorship and peer learning opportunities are provided for continuous support. Practical tools are shared to equip healthcare workers with necessary skills. Cultural sensitivity is fostered to ensure healthcare workers can provide care that is respectful and responsive to diverse cultural backgrounds and language preferences. By incorporating these strategies, the training program aims to empower healthcare workers with the expertise and empathy needed to provide inclusive and effective healthcare services for persons with disabilities.

## Discussion

Developing a comprehensive training program for healthcare workers on disability is an important step in addressing barriers to healthcare access for people with disabilities. We used the Medical Research Council approach to develop the training material (16), which considering practical solutions to improve the knowledge and skills of healthcare workers on disability. Our umbrella literature search revealed that lectures and didactic methods were commonly used in disability education, often combined with other approaches such as case studies and simulations. The review also highlighted the importance of using various teaching methods and including people with disabilities in disability education. The impact of education was measured in various ways, through healthcare worker reports on comfort, attitudes, communication skills, knowledge, and self-efficacy. There is need for a standardised approach to allow comparison between contexts and countries. Standardisation in curricula and limited time and resources were identified as challenges in providing disability education within healthcare training.

People with disabilities and healthcare workers in Uganda expressed the need for better communication skills from healthcare providers, training on disability, and recognition of challenges faced by people with disabilities. These findings are echoed in other studies globally (18–21). The workshop successfully generated a comprehensive and inclusive framework for healthcare worker training on disability, incorporating the diverse perspectives of people with disabilities and healthcare workers. Our findings are consistent with other studies that highlight the importance of disability education in healthcare training, utilising various teaching methods and incorporating the perspectives of people with disabilities (22, 23). The next step is to pilot-test the training programme with healthcare workers in Luuka District, Uganda (planned October, 2023). During pilot testing, the training program will also consider the cultural context in which it will be delivered, as previous research has shown that cultural competence is essential for providing effective and appropriate care to people with disabilities (24, 25).

One of the strengths of our study was the use of the MRC approach to develop the training material, which enabled us to systematically examine training needs and develop practical solutions to improve the knowledge and skills of healthcare workers on disability. However, a limitation is that it only represents the perspectives of people with disabilities and healthcare workers in Uganda, and thus its generalisability to other contexts may be limited. While we did include the opinions of international experts and a global umbrella review, further research is needed to confirm the effectiveness and applicability of the training program in different settings, as the umbrella review noted important gaps in the literature.

Our findings have important implications for policy and programmes and research. Specifically, our study suggests that healthcare worker training on disability should be inclusive of diverse cultural backgrounds, flexible and adaptable to specific needs, and should incorporate the perspectives of both people with disabilities and healthcare workers. These findings could be used to inform the adaptation of future training programs for healthcare workers in Uganda and other resource-limited settings. Policy support is vital to ensure implementation and support of training of healthcare workers on disability (e.g. mandating inclusion in medical and nursing curricula). Future studies could use more objective measures of impact, such as patient outcomes or changes in healthcare service provision.

## Conclusion

The proposed development of training for healthcare workers on disability aims to address the barriers and challenges faced by people with disabilities in accessing health care services in Africa. The training will adopt a modular approach and will be based on the African health disability perspective. The most important components of the training include the emphasis on adopting a disability-inclusive approach, interaction with people with disabilities, a learner-centred and participatory approach, ongoing mentorship and peer learning, practical tools to deliver a comprehensive and considerate examination and treatment, and cultural sensitivity.

## Declarations

### 1. Ethics declarations and consent to participate

All protocols were approved by the Uganda Virus Research Institute research ethics committee (ref: GC/127/904) and the London School of Hygiene & Tropical Medicine (ref: 26715) and the Uganda National Council of Science and Technology (SS1348ES).This study was conducted in accordance with the Declaration of Helsinki. Informed consent was obtained from all participants. Written informed consent was obtained from participants (people with disabilities and healthcare workers) who undertook interviews that were conducted in Uganda. Verbal informed consent was obtained from stakeholders, whose interviews were online, and the ethics committee approved this procedure.

### 2. Consent for publication

Participants consented for publication of anonymised quotes

### 3. Availability of data and materials

The majority of the datasets supporting the conclusions of this article are included within the article and its additional files. However, underlying interview data associated with this study will not be made freely available, as the small number of healthcare worker and people with disabilities makes data potentially identifying. Applications for access to the raw qualitative data for this study should be made via email to the corresponding author, outlining the purpose of the proposed analyses and the data requested. These applications will be reviewed by the LSHTM’s data access committee, and if accepted, the requested data will be shared.

### 6. Author contributions

Conception or design of the work: TS, HK

Data collection: TS, ASS, SS

Data analysis and interpretation; TS, ASS, SS

Drafting the article: TS

Critical revision of the article: TS, ASS, SS, FBM, HK

Final approval of the version to be submitted - all named authors

## 7 Acknowledgements

Not applicable

